# Use of oral Semaglutide after Acute Coronary Syndrome - Design and rationale of a prospective observational study

**DOI:** 10.1101/2025.08.03.25332910

**Authors:** M. Biasin

## Abstract

**Rationale:** Oral semaglutide, a glucagon-like peptide-1 receptor agonist (GLP-1 RA), has shown cardiovascular benefits in patients with type 2 diabetes mellitus (T2DM) and established atherosclerotic cardiovascular disease (ASCVD). However, its initiation in patients immediately following an acute coronary syndrome (ACS) has not been prospectively investigated. This study addresses this critical gap by evaluating the feasibility, tolerability, and clinical characteristics associated with prescribing oral semaglutide (Rybelsus) at hospital discharge in patients with T2DM admitted for ACS.

**Objectives:** The primary objective is to evaluate the proportion of patients discharged after ACS who successfully initiate and tolerate oral semaglutide within 30 days, without permanent discontinuation due to adverse events by 3 months. Secondary objectives include characterization and quantification of adverse events (with a focus on gastrointestinal intolerance, pancreatitis, and worsening renal function), assessment of treatment persistence and adherence, evaluation of metabolic parameter changes, documentation of cardiovascular outcomes during 12 months of follow-up, and exploration of clinical predictors for treatment discontinuation and adverse events.

**Methods:** This is a prospective, observational, multicenter study enrolling adult patients with T2DM admitted for ACS (STEMI, NSTEMI, or unstable angina), discharged from the coronary care unit with documented initiation of oral semaglutide (Rybelsus). Patient data will be systematically collected at baseline (hospital discharge), 3, 6, and 12 months, through standardized electronic case report forms during clinical visits and via electronic health records. Collected data includes demographics, ACS clinical presentation, metabolic parameters (HbA1c, BMI, eGFR), semaglutide dosing, adherence, adverse events, treatment continuation rates, and cardiovascular outcomes.

**Ethics and Dissemination:** Ethical approval will be obtained from institutional review boards in accordance with national regulations. All participants will provide informed consent. Findings will be disseminated through peer-reviewed publications and presentations at scientific conferences.

**Trial Registration:** Not applicable. This is an observational, non-interventional study

## Background and Rationale

Oral Semaglutide, a GLP-1 receptor agonist, has demonstrated cardiovascular benefit in patients with type 2 diabetes mellitus (T2DM) and established atherosclerotic cardiovascular disease (ASCVD) but its role in the immediate post-acute coronary syndrome (ACS) setting remains underexplored. As of now, there are no studies published on PubMed specifically investigating the prescription or initiation of oral semaglutide at hospital discharge following an ACS event.

Available evidence from the PIONEER program has shown the cardiovascular safety and metabolic efficacy of oral semaglutide in patients with T2DM, but these trials enrolled stable patients rather than those recently discharged post-ACS^1^. The SOUL trial, published in 2025, demonstrated a reduction in major adverse cardiovascular events (MACE) with oral semaglutide in patients with T2DM and ASCVD, but again, it did not include patients treated at the time of discharge after ACS^2^. Additionally, a recent Italian study by De Sio et al. evaluated the eligibility of overweight or obese post-ACS patients without diabetes for GLP-1 RA therapy but did not assess the timing of initiation or specifically focus on Rybelsus^3^.

Given the known cardiovascular benefits of GLP-1 RAs and their growing role in cardiometabolic management, this study aims to address a gap in the literature by prospectively evaluating the feasibility, safety, and clinical characteristics of patients discharged from the coronary care unit (CCU) who were prescribed Rybelsus. By evaluating real-world initiation of oral semaglutide in the immediate post-ACS setting, this study may provide novel insights into early cardiometabolic optimization in high-risk patients.

### Study Design

- Prospective, multicenter, observational study.
- Study period: January 2026 to December 2026.
- Patients will be enrolled at hospital discharge following ACS.
- Data will be collected at baseline (discharge), 3, 6, and 12 months via clinical visits and/or electronic health records.
- Data collection will be harmonized across centers using a standardized electronic case report form

## Study Objectives

### Primary objective

- **To evaluate the proportion of patients discharged after ACS who successfully initiate and tolerate oral semaglutide within 30 days**, without permanent discontinuation due to adverse events by 3 months.

### Secondary objectives

1. **To quantify the prevalence and type of adverse events (AEs)** possibly related to semaglutide at 3, 6, and 12 months, with particular focus on:
  ∘ Gastrointestinal intolerance (nausea, vomiting, diarrhea, early satiety)
  ∘ Pancreatitis (clinically suspected or confirmed)
  ∘ Worsening renal function (≥30% increase in serum creatinine from baseline)
2. **To assess treatment persistence and adherence**:
  ∘ Proportion of patients continuing semaglutide at each follow-up (3, 6, and 12 months)
  ∘ Proportion of patients reaching or maintaining the 14 mg daily dose
3. **To evaluate changes in metabolic parameters** between baseline and follow-up:
  ∘ HbA1c variation
  ∘ Body weight and BMI variation
  ∘ Changes in eGFR
4. **To assess cardiovascular outcomes over 12 months**, including:
  ∘ All-cause mortality
  ∘ Cardiovascular death
  ∘ Non-fatal myocardial infarction
  ∘ Non-fatal stroke
  ∘ Unplanned coronary revascularization
  ∘ Hospitalization for heart failure
5. **To explore predictors of treatment discontinuation and adverse events**, including age, renal function, baseline HbA1c, BMI, and type of ACS.

### Study Population

**Inclusion** criteria:

- Age ≥18 years.
- Hospital admission for ACS (STEMI, NSTEMI, or unstable angina).
- Diagnosis of type 2 diabetes mellitus.
- Discharged from ICCU with a documented recommendation to initiate **oral semaglutide** (Rybelsus).
- Willingness and ability to attend scheduled follow-up visits for at least 12 months. **Exclusion** criteria:
- End-stage renal disease (eGFR <15 mL/min/1.73m^2^).
- Terminal illness with life expectancy <6 months.
- Contraindications to GLP-1 receptor agonists.

### Data Collection

Data will be collected at four pre-specified timepoints:

- **Baseline** (at hospital discharge)
- **3 months, 6 months**, and **12 months** after discharge.

**Variables collected at each visit:**

| TIMEPOINT | VARIABLES |
| --- | --- |
| **BASELINE** | Age, sex, BMI, T2DM history, HbA1c, systolic/diastolic BP, lipid profile, eGFR, ACS type, Killip class, LVEF, PCI/CABG, medications (including GLP-1 RA initiation) |
| **3 MONTHS** | Weight, HbA1c, eGFR, adherence to Rybelsus, adverse events, hospitalization, CV events |
| **6 MONTHS** | Same as 3-month, with confirmation of treatment continuation/discontinuation |
| **12 MONTHS** | Same as above; documentation of vital status, long-term adherence, final outcome |

In addition to structured visits, unscheduled events (e.g., CV readmission, adverse events) will also be documented when applicable.

### Statistical Analysis

- Descriptive statistics will be used to summarize baseline demographics, comorbidities, ACS characteristics, and treatment data. Continuous variables will be reported as mean ± SD or median [IQR], as appropriate; categorical variables will be reported as counts and percentages.
- Kaplan–Meier survival analysis will be performed to estimate time to major adverse cardiovascular events (MACE) and treatment discontinuation, with log-rank tests for group comparisons.
- Cox proportional hazards models will be used to identify independent predictors of MACE and discontinuation, adjusting for clinically relevant covariates (e.g., age, sex, eGFR, HbA1c, ACS type).
- Logistic regression will be applied for binary outcomes at specific timepoints (e.g., occurrence of adverse events at 3 or 6 months).
- Linear mixed-effects models or generalized estimating equations (GEE) will be used to analyze changes in repeated measures (HbA1c, weight, eGFR) over time, accounting for intra-subject correlation and missing data.A per-protocol sensitivity analysis will be performed to evaluate outcomes among patients who reach or maintain the target dose (14 mg/day

### Ethical Considerations

- The protocol will be submitted to the ethics committees of participating centers.
- Informed consent will be obtained from all patients prior to enrollment.
- Data will be pseudonymized and handled in accordance with GDPR and national privacy laws.

## Data Availability

All data produced in the present study are available upon reasonable request to the authors

